# Withdrawal of Assisted Ventilation at the Patient’s Request in MND/ALS: A Retrospective Exploration of the Ethical and Legal Issues Concerning Relatives, Nurses and Allied Health Care Professionals

**DOI:** 10.1101/2022.03.14.22271768

**Authors:** K. Phelps, E. Regen, C.J. McDermott, D.J. Oliver, C. Faull

**Affiliations:** Department of Health Sciences, University of Leicester, Leicester, UK; Department of neurology, Sheffield Institute for Translational neuroscience, University of Sheffield, Sheffield, UK; Tizard Centre, University of Kent, Canterbury, UK; LOROS: The Leicestershire and Rutland Hospice, Leicester, UK; Honorary Professor, Department of Health Sciences, University of Leicester, Leicester UK; University Hospitals of Leicester, Leicester, UK

**Author notes:** **Correspondence** Professor C Faull, LOROS, Groby Road, Leicester LE3 9QE.

**Keywords:** Motor Neurone Disease (MND), MND, Amyotrophic lateral sclerosis (ALS), Ventilation, Ethics, Legal, Withdrawal of treatment, Nurses, Allied health professionals, Family

## Abstract

**Background:** There is little literature focusing on the issues relatives and health professionals encounter when withdrawing assisted ventilation at the request of a patient with MND/ALS.

**Aim:** To explore with relatives, nurses and allied health professionals the ethical and legal issues that they had encountered in the withdrawal of ventilation at the request of a patient with MND/ALS.

**Method:** A retrospective qualitative interview study with 16 family members and 26 professionals. Data was analysed thematically and compared with results from a previous study with doctors.

**Results:** The events surrounding ventilation withdrawal were extraordinarily memorable for both HCPs and family members with clear recall of explicit details, even from years previously. The events had had a profound and lasting effect due to the emotional intensity of the experiences. Withdrawal of ventilation posed legal, ethical and moral challenges for relatives and health are professionals. Relatives looked to health care professionals for knowledge, guidance and reassurance on these issues, worried about how the withdrawal would be perceived by others, and found professional ignorance and disagreement distressing. Many health care professionals lacked theoretical knowledge and confidence on the legal and ethical considerations of withdrawal and struggled morally knowing the outcome of the withdrawal would be death. Health care professionals also worried about the perception of others of their involvement, which in turn influenced their practice. There was a lack of consistency in understanding across professions, and professionals often felt uncomfortable and anxious

**Conclusions:** Legal, ethical and practical guidance is needed and open discussion of the ethical challenges as well as education and support for health care professionals and relatives would improve the experience of all involved.

## BACKGROUND

Motor neurone disease or amyotrophic lateral sclerosis (MND/ALS) is a progressive neurological condition which primarily affects motor neurons leading to skeletal muscle weakness. Most deaths in MND/ALS are due to respiratory failure, as a result of diaphragm and respiratory muscle weakness.^1^ Over the last 10 years non-invasive ventilation (NIV) has been increasingly used and it has been shown to support ventilation very effectively, often for many months, improving quality of life, symptoms and survival.^2-4^ Neurological deterioration is relentless for patients despite ventilation and patients may eventually reach a point at which they cannot move or communicate - becoming ‘locked in’.^5^ Many patients also develop cognitive change which can affect their ability to make decisions.^6^ There is therefore considerable need to prepare patients for their deteriorating condition and allow them to be involved in decision making, ahead of the situation occurring.^3,7,8^

Studies have shown that fear of dependence on ventilation is a common negative experience for people when starting on NIV.^9^ Many patients with MND/ALS who use NIV in the UK either stop the intervention themselves as they deteriorate or die whilst still using it.^10^ However some patients who are dependent on NIV, and some on Trachea Ventilation (TV), do wish to stop because they can no longer tolerate it or because of deterioration in other factors impacting on their quality of life. Some may make a written statement with respect to withdrawal in advance of their losing the ability to communicate or other reason for loss of capacity.^11^

A previous study identified the significant challenges palliative medicine doctors in the UK experienced in withdrawal of assisted ventilation at the request of a patient with MND/ALS.^12^ We explored these in more depth with a broader group of doctors and have reported significant ethical and legal challenges that doctors have experienced.^13^ This paper reports on the experiences of family members, nurses and allied Health Care Professionals (HCPs) who have been involved with withdrawal of ventilation at the request of a patient. The data collection was undertaken in 2014, prior to development of professional Guidance^14^.

## METHODS

This was a retrospective qualitative interview study. Family members were asked to discuss their experience of their loved one with MND/ALS who had requested withdrawal of ventilation. HCPs were asked to talk about their experiences of MND/ALS patients with whom, at the request of the patient, they had been involved in withdrawal of ventilation. A topic guide, developed from previous studies,^13^ was utilised flexibly to explore experiences in detail with heath care professionals. Interviews with family members were based on them telling their story, whilst picking up and exploring in detail the lead up to, and carrying out the ventilation withdrawal.

### Participants and data collection

HCPs, across England and Wales, who had been involved in withdrawal of ventilation at the request of a patient with MND/ALS within the past 5 years, were invited to participate in a one to one interview in person or by telephone, as they preferred. A multifaceted recruitment strategy was employed to approach participants as described in a previous report.^13^

Family members of people who had been withdrawn from NIV were identified by clinicians in our previous study, HCPs in the current study or through the MND Association. They were provided with information sheets about the project and those who were interested in taking part contacted the research team to discuss the study and arrange a time and place for interview.

#### Participants: Health care professionals

Twenty six nurses and allied health care professionals (7 palliative care, 10 ventilation/ respiratory, 6 neurology/ MND/ALS and 3 community nurses) (see table 1) were interviewed, 2 by telephone and 24 face to face. Community nurses and those specialising in palliative care were likely to have been involved in only one withdrawal. Those specialising in ventilation/respiratory care and Neurology/ MND/ALS were more likely to have experienced more than one withdrawal, but none had been involved in more than 3 cases. In total thirty five cases of ventilation withdrawal from patients with MND/ALS were discussed, some being discussed by more than one professional, and some professionals discussing more than one case.

**Table 1.** HCP Participants Roles and Experience.

| **Code** | **Role** | **Cases discussed during interview** |
| --- | --- | --- |
| HCP01 | Clinical Nurse<br>Specialist Palliative<br>Care | 2 cases of NIV withdrawal at home<br>NB: Interviewee refused to be involved in actual<br>withdrawals |
| HCP02 | Nurse Palliative<br>Care | 2 cases of NIV withdrawal at home (male) – see also<br>R02 & R03 |
| HCP03 | Specialist<br>Neuromuscular<br>Respiratory<br>Physiologist | 1 case of NIV withdrawal at a hospice<br>1 case of NIV withdrawal at home – see also R04 |
| HCP04 | Clinical Nurse<br>Specialist Palliative<br>Care | 1 case of NIV withdrawal at home– see also HCP15 |
| HCP05 | Senior<br>Physiotherapist<br>Palliative Care | 1 case of NIV withdrawal in hospice |
| HCP06 | Clinical Nurse<br>Specialist sleep<br>disorders | 1 case of IV withdrawal at home<br>1 case of NIV withdrawal in a nursing home |
| HCP07 | Lead Nurse<br>Ventilation | 1 case of NIV withdrawal at home– see also R06 |
| HCP08 | Clinical<br>Physiologist<br>Ventilation | 2 cases of NIV withdrawal at home |
| HCP09 | Clinical Nurse<br>Specialist<br>Ventilation | 1 case of NIV withdrawal at home– see also R11,<br>HCP17, HCP20 |
| HCP10 | Clinical Nurse Specialist<br>Neurology Rehab | 3 cases of NIV withdrawal in hospital |
| HCP11 | Nurse Consultant<br>Home Ventilation<br>Service | 1 case of IV withdrawal at home – see also R07, HCP19<br>1 case of IV withdrawal in hospital– see also HCP19 |
| HCP12 | Specialist<br>Respiratory Nurse | 1 case of NIV withdrawal in hospital– see also HCP 14<br>1 case of NIV withdrawal at home– see also R09,<br>HCP14,<br>1 case of IV withdrawal in hospital |
| HCP13 | Community Nurse | 1 case of NIV withdrawal at hospice – see also R01, |
| HCP14 | MND Specialist<br>Nurse | 1 case of NIV withdrawal at home– see also R09,<br>HCP12,<br>1 case of NIV withdrawal in hospital – see also R08, |
| HCP15 | Community Nurse | 1 case of NIV withdrawal at home– see also HCP04 |
| HCP16 | Community<br>Palliative Care<br>Specialist Nurse | 1 case of NIV withdrawal at hospice |
| HCP17 | Community<br>Specialist Palliative<br>Care Nurse | 1 case of NIV withdrawal at home– see also R11,<br>HCP09, HCP20 |
| HCP18 | Senior<br>Physiotherapist<br>Palliative Care | 1 case of IV withdrawal at home – see also HCP24,<br>2 cases of NIV withdrawal<br>at home see also HCP24, |
| HCP19 | Respiratory Nurse | 1 case of IV withdrawal at home– see also R07, HCP11,<br>1 case of IV withdrawal in hospital– see also HCP11 |
| HCP20 | District Nurse<br>Team Leader | 1 case of NIV withdrawal at home– see also R11,<br>HCP09, HCP17 |
| HCP21a | Social Worker | 1 case of IV withdrawal at home– see also R10 |
| HCP21b | MNDA visitor |  |
| HCP22 | Specialist MND Nurse | 1 case of NIV withdrawal at home<br>2 cases of NIV withdrawal in hospital ( |
| HCP23 | MND Regional Care Development Advisor | 1 case of IV withdrawal see also R05 |
| HCP24 | Community Palliative care Nurse | 1 case of IV withdrawal at home – see also HCP18,<br>1 case of NIV withdrawal at home– see also HCP18 |
| HCP25 | Ventilation Specialist Nurse | 1 case of IV withdrawal at home– see also R16, |

Sixteen HCP’s had been involved with NIV withdrawal(s) only, four had been involved with TV withdrawal(s) only, and four had been involved in both NIV and TV withdrawals. Thirteen respondents had been involved with more than one patient withdrawal. The settings for the withdrawal included home (most common for TV), hospice, nursing home and hospital.

#### Participants: Relatives

Sixteen family members of MND/ALS patients who had withdrawn from ventilation took part in face to face interviews. Of these, fourteen were spouses who were interviewed alone and two were a spouse and adult child who were interviewed together. The characteristics of the person who had died are shown in table 2. Two were female and fourteen male, twelve had withdrawn from NIV and four from TV. The setting for the withdrawal had been at home (twelve cases), hospice (three cases) and hospital (one case).

**Table 2.** The context of the person with MND who had stopped their ventilation support spoken about by the relative participants.

| **Relative participant** | **Age and gender of person with MND** | **Type of ventilation withdrawn** | **Place of withdrawal** | **Case also discussed by:** |
| --- | --- | --- | --- | --- |
| R01 | Female<br>56-65 | NIV | Hospice | HCP13 |
| R02 | Male<br>56-65 | NIV | Home | HCP02 |
| R03 | Male<br>56-65 | NIV | Home | HCP02 |
| R04 | Female<br>66-75 | NIV | Home | HCP03 |
| R05 | Male<br>66-75 | TV | Home. | HCP23 |
| R06 | Male<br>56-65 | NIV | Home | HCP07 |
| R07 | Male<br>46-55 | TV | Home | HCP11,<br>HCP19 |
| R08 | Male<br>56-65 | NIV | Hospital | HCP14 |
| R09 | Male<br>66-75 | NIV | Home | HCP12,<br>HCP14 |
| R10 | Male<br>56-65 | TV | Home | HCP21 |
| R11 | Male<br>36-45 | NIV | Home | HCP09,<br>HCP17,<br>HCP20 |
| R12 | Male<br>56-65 | NIV | Home |  |
| R13 | Male<br>56-65 | NIV | Home |  |
| R14 | Male<br>56-65 | NIV | Hospice |  |
| R15 | Male<br>56-65 | NIV | Hospice |  |
| R16 | Male<br>46-65 | TV | Home | HCP25 |

### Data Analysis

The interviews were audio-recorded and transcribed and interviewers (ER & KP) maintained reflective diaries. Data from transcripts were analysed by constant comparison based on grounded theory to identify themes,^15,16^ as described in a previous paper.^13^

### Ethical concerns

The study was approved by the East Midlands Derby Research Ethics Committee - reference 11/EM/0131. All participants were provided with participant information sheet that explained their involvement in the study.

## RESULTS

The events surrounding the ventilation withdrawal were extraordinarily memorable for both HCPs and family members with clear recall of explicit details, even from years previously. The events had had a profound and lasting effect due to the emotional intensity of the experiences. This paper focuses on HCP and relatives’ experiences of the ethical and legal issues around withdrawal of ventilation at the request of the patient with MND/ALS. The impact of HCPs ethical and legal concerns on patient care and family experience is highlighted. Other themes are discussed in separate papers.

### Lack of knowledge of the Legality and Ethics of Withdrawal of Ventilation

Most relatives and many HCPs talked of their initial belief that removal of ventilation was assisted suicide or euthanasia. The level of understanding of the theory was greater amongst those who specialised in ventilation or had previous experience of withdrawal. The community nurses in our sample seemed unsure of the ethics of withdrawal prior to more detailed discussions with colleagues.

Legal and ethical misconceptions seemed to arise from the perception that removal of ventilation causes the death. Those with experience of ventilation recognised that death from MND/ALS could only be postponed by ventilation. HCPs with experience of patients dying whilst still on ventilation were clearer about the ethical and legal issues. The lack of written professional guidance or literature on the subject meant that HCPs and relatives were dependent on the knowledge of those around them, which in many cases was confused and sometimes inaccurate. Confusion and lack of knowledge among HCPs could be perceived by patients and their families, and added to their anxieties at a very stressful time.

> *HCP13 Ethically I think it felt OK because it was withdrawing an intervention that had been added in. And I think if you’re giving medication to relieve panic and distress, then that’s not the same as giving medication to finish somebody off. (Community Nurse)*

#### Fear of being viewed by others as assisting suicide

Many of the HCPs in our study were afraid of being accused of having assisted a patient to die. Respondents were concerned that colleagues around them would think they had done something unethical. There was a fear of possible reprisals stemming from these misunderstandings, such as court cases and disciplinary action at work. Some HCPs did feel that their level of professional protection and support was not as great as that of the doctors and felt quite vulnerable. Table 3 identifies the difficulties that had been encountered by participants.

**Table 3.**
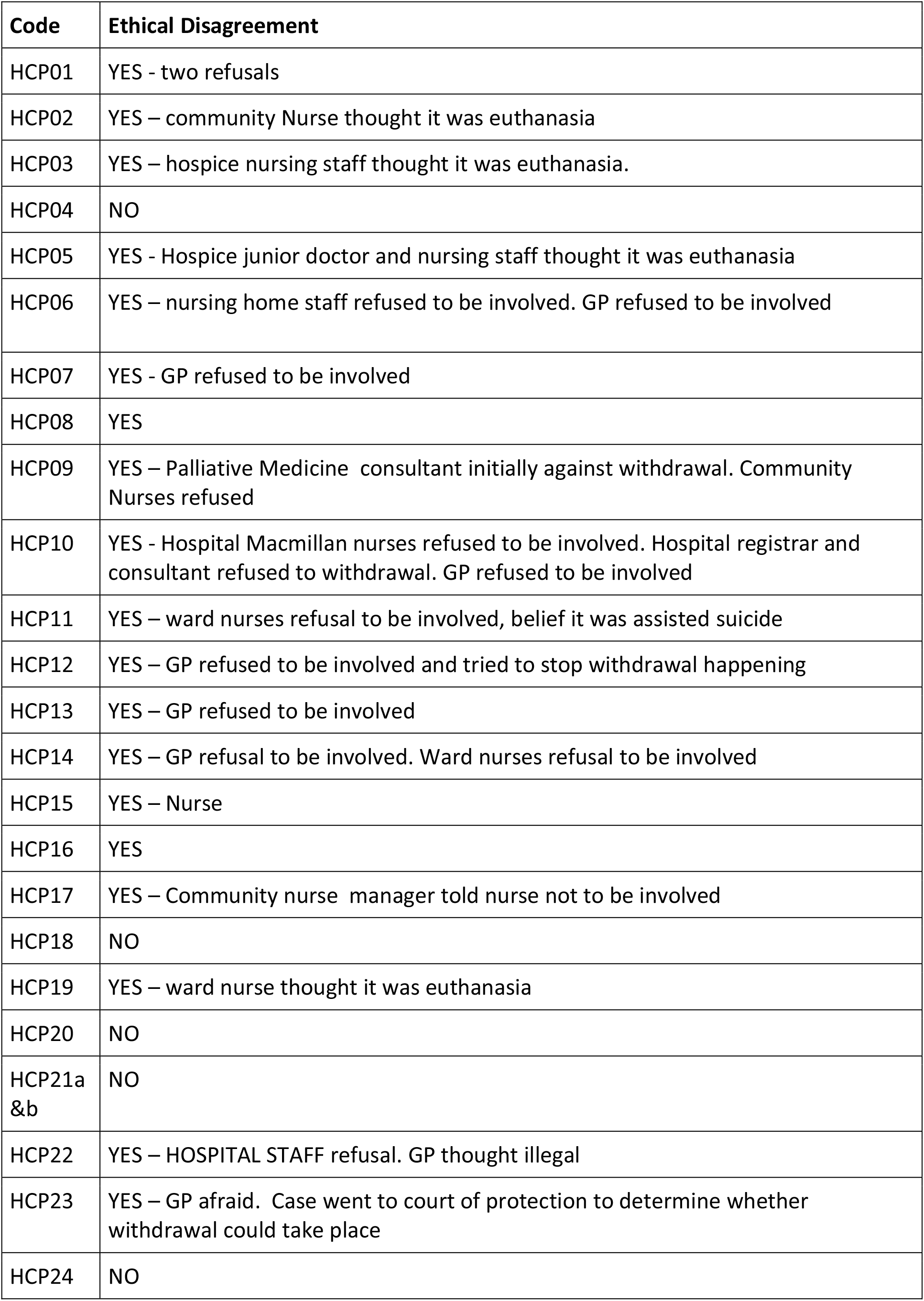
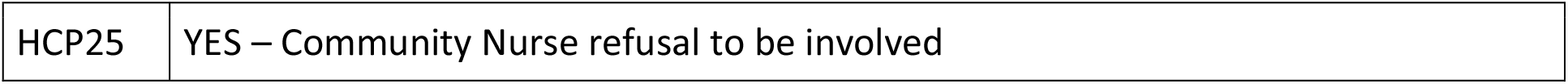
Health professional experience of disagreement between professionals about the ethics and legality of withdrawal and the impact on care.

Many of our respondents spoke of being afraid of the press misrepresenting cases of assisted ventilation withdrawal, and making it seem like assisted suicide

> *HCP04 … if that story had been in the news, how would that have been perceived? And would it be the case that some local newspaper would then suddenly say ‘palliative care nurses can now help you die’. And you know and if it’s a tabloid they’re not going to want to engage in the ethics of it, yeah. So I think that was something that was on my mind a little bit. (Palliative specialist)*

Many family members were also afraid that they would be viewed by others as having helped their loved one to die or not prevented it. They often did not tell friends and family that the patient had chosen to withdrawal from treatment for fear of the reaction and lack of understanding of the legal and ethical position. In more than one case there were other family members or staff who did feel that the withdrawal was assisted suicide or murder which family members found deeply upsetting and it had a lasting impact on their memories of their loved one’s death.

#### The impact of the link between withdrawal and patients expressed wish to die

The ethical and legal complexity for relatives and HCPs was confounded if it seemed that the underlying reason for the patient’s request to withdraw from treatment was their wish to die rather than the removal of a burdensome treatment. This made the process seem more like assisted suicide, rather than treatment withdrawal. HCPs spoke of patients having ‘had enough’ and their quality of life having deteriorated to such an extent that they found continued living, made possible by the assisted ventilation, unacceptable. The impact of this was twofold. In some cases it precipitated a conversation about withdrawal of ventilation, however it also led to a cognitive link between the patients request to die and the withdrawal.

Many family members stated that the patient had at some point come to the stage in their illness when they no longer wished to be kept alive, and at this point had made a request for assistance to die. These requests were sometimes made to the family member and sometimes to a doctor or other HCP. Relatives felt distressed and sometimes pressured by their loved one’s request, and some felt morally obligated to try and assist them to die although they were fearful of the consequences for themselves. Requests to doctors and HCPs for assistance to die were met with resistance (due to the illegality of the act in the UK), but often precipitated a discussion about the patient’s right to choose to stop using assisted ventilation and the alternative support that professionals could provide to manage symptoms.

HCPs reluctance to discuss withdrawal due to their lack of understanding and subsequent fear meant that there were many cases where the patient and family were unaware of the right to withdraw treatment until the patient expressed a desire to die. Family members felt that if they had known earlier that it was legal to withdraw, they could have been spared much anguish. Some relatives had experienced considerable agonising over how to help their loved one, relieve their suffering and carry out their wishes. Many respondents felt that if patients had been made aware of their right to withdraw treatment at an earlier stage then they may have requested it sooner and suffered less.

> *R13 … he went back in hospice again just for a week not long before he died and that’s when he come to a decision that enough were enough and we had a real, some really good chats with the doctor and he said that it were possible that they could make him comfortable. The role they could play was making him comfortable to be able to come off the machine. Obviously it was his decision. And he said that’s what he wanted to do*.
>
> *R14 Until he said he didn’t want to carry on. That it wasn’t right that there wasn’t euthanasia here, and he was quite adamant that people should have that choice. It was then that they said about this [withdrawal] but it had never been mentioned before*.

#### The need for an ethical and legal framework for ventilation withdrawal in MND/ALS

The majority of HCPs were of the view that clear guidance would be very helpful for professionals struggling with the ethical and legal issues surrounding withdrawal of ventilation, and could relieve some of the ethical burden and emotional stress they faced. They felt that they would have been reassured by a clear statement by a professional body, stating that withdrawal of ventilation is legal and ethical and under what circumstances. Respondents felt that professional guidelines would give professionals something to show colleagues who were misinterpreting the withdrawal as assisted suicide and reduce some of the fear of reprisals. Family members who felt that clarity would improve the care received by patients dying from MND/ALS, and reduce the burden on their families echoed this view.

### The impact for patients and families on the practical aspects of withdrawal

Undertaking the withdrawal from ventilation was complex, and practice varied considerably between cases. This was due in part to the legal, moral and ethical uncertainty of those involved.

#### Experience Matters

This is a situation where patients and their families need the HCPs around them to be confident and sure of what they are doing. The family members, whose medical team was experienced, felt that the process was much smoother and less traumatic than for those whose team were inexperienced. If patients and their families received unsure or conflicting information from HCPs then this added to their (already considerable) distress, level of fear and weight of responsibility. For those families whose medical team was in agreement and confident about the ethics and legality of withdrawal, these fears and anxieties were greatly lessened.

All of the HCPs in our study had discussed the ethics and legality of their cases with colleagues and clinicians, both within their teams and outside. Those with experience provided advice to others and those without sought clarification of the issues and legal standpoint. For many there was a strong need for support and advice with ethical and legal issues, and considerable anxiety caused by the fear of getting it wrong. There was a clear view that HCPs should not be dealing with this situation alone, and some kind of formal supervision should be available. There was also feeling that HCPs need the support of a doctor if they are to be involved. Those who did not have the backing of a MDT could feel very vulnerable and in fear of repercussions.

Two of the HCPs felt the need to consult more formally over ethics and the law, with their employer’s legal department or professional body, but the majority of HCPs did not feel it necessary to obtain formal advice or support. Either because the felt confident of the legal position or because they were working with a clinician who took that responsibility.

All of the family members in our study had discussions with at least one HCP about the ethics and legality of withdrawal of ventilation. Most wanted reassurance that this was a legal and ethical procedure, and not assisted suicide. In some cases relatives and patients spoke to religious leaders for advice. In many cases the ethical and legal position needed to be explained to many family members, not just the patient and their main carer. This could be the patient’s children, brothers and sisters, or even parents. HCPs and family members wanted, if possible, to ensure all were in agreement and there were no misunderstandings about the patient’s cause of death. The importance of HCPs having full understanding of ethical and legal concerns so that they can make the position clear to patient and families cannot be overstated.

#### The impact of ethical and legal disagreement on patient care

All but one of the HCPs in our study were prepared to support their patients during withdrawal. The impact on patients and families if HCPs refuse to be involved in withdrawal was potentially substantial, and could impact on place of death and care received. One of our respondents had twice declined to be involved in ventilation withdrawal due to her personal belief that it was unethical and immoral to do so. Although she understood quite clearly the legal standpoint, she still felt it to be wrong.

Experience of professional disagreement as to whether withdrawal of ventilation was ethical and legal was common in our sample. Twenty of our twenty-six HCPs had experienced other health care professionals refusing to take part in a ventilation withdrawal and/or believing it to be illegal or unethical. These included hospital nursing staff, hospice nursing staff, community nurses, GPs and a hospice doctor. Six of the cases discussed by our respondents involved the refusal of GPs. The effect this had on care for patients varied considerably, depending on the other staff involved in the cases. Impacts for patients and families included long delays in withdrawal taking place, patients not being able to die in their preferred place, unfamiliar staff being involved, and lack of basic end of life care such as symptom control. When patients and families were aware of these refusals there was considerable distress.

> *HCP14 … went to speak to the GP who was, I think I would describe it – his response – as a – even over the telephone – as a rabbit in headlights. Absolutely ‘I’m not sure about this. Is this legal?*… *(MND/ALS specialist)*
>
> *HCP07 She (GP) said she was going to have absolutely nothing to do with removing the ventilator or touching the settings or…which made it very difficult because I couldn’t guarantee that I was going to be available around the clock to do it, whereas she could have been, more so, or she could have, perhaps, interacted with me. (Ventilation specialist)*

Some relatives had experienced disagreement from HCPs and professional carers who were uncomfortable with the prospect of withdrawal or even felt withdrawal to be assisted suicide or killing. Some relatives felt that this was to do with religious beliefs. One relative was very distressed at being accused by one of the patient’s carers of ‘coercing’ the patient into withdrawal, an accusation she found deeply offensive. In this specific case the articulation of the carer’s view lead to the withdrawal being delayed for many months causing considerable suffering for the patient and their family.

#### Protecting the family from the burden of guilt and fear of repercussions

There were many examples recalled by family participants where patients and families were spared some of the moral and ethical burden and fear of legal repercussions from the withdrawal by the good practice of the health care professionals around them.

□ Where patients and families were made aware of the patients legal right to withdrawal from treatment before the patient lost communication or progressed to an advanced stage of the disease
□ Where clear, unambiguous, correct statements of the ethics and legality of ventilation withdraw had been given to patients and their families
□ Where HCPs assisted in family disagreements by explaining the ethical and legal position to family members and protected them from accusations of assisted suicide
□ Where relatives and patients were kept unaware of the ethical and legal disagreements and discussions going on within the medical team
□ Where the withdrawal was carried out by a professional team with no need for relatives to take part, except to be with their loved one as they withdrew and died.

There were also examples where patients and families could have been spared some of the moral and ethical burden and fear of legal repercussions from the withdrawal. Patients and families, already in distress, experienced greater emotional upset by the action or inaction of the HCPs involved

□ Where patients and families were not made aware of the patients legal right to withdrawal from treatment until such time as the patient requested help to die
□ Where patients and families were given incorrect or conflicting ethical / legal advice
□ Where patients and families were refused withdrawal from a medical professional, or offered it reluctantly with strings attached
□ Where patients and families were aware of disputes within the medical team
□ Where families received little support for the withdrawal or were left to carry out the withdrawal themselves and were left with feelings of guilt and doubt

#### Impact of ethical-legal issues on practice

Many HCPs felt the need to keep a detailed written account of the withdrawal in case any questions were asked at a later date about the ethics and legality of their practice. The detailed records kept in these cases were over and above ‘usual practice’, undoubtedly due to the fear of possible ethical and legal repercussions. In some cases this was carried out in a group and those present all signed the written report of the withdrawal. Information documented in detail included:

□ Discussions prior to withdrawal, who was involved, and what was discussed
□ Who was involved in the withdrawal and who did what
□ Symptom control medication used during withdrawal, administration and timing
□ Ventilator levels, how much they were reduced by, and how often if weaning was used.

There was some feeling that a doctor should be present at a withdrawal to carry the ethical and legal responsibility for the withdrawal, and in particular to manage the medication used to manage symptoms. Some HCPs had experience of clinicians who insisted on taking the responsibility so as to protect other professionals involved from possible repercussions.

> *HCP15 … but the GP said “I’m here. Whilst I’m here I’ll do it because that’s not going on your shoulders” basically. (Community nurse)*

More than one of our respondents faced external scrutiny of their practice after a withdrawal, voluntarily or due to questions being raised about ethical practice. One group of HCPs who had experience of 2 withdrawals in which there was professional disagreement chose to discuss these cases at their hospital ethical committee. They were anticipating future cases where this may occur again and felt that backup from the committee would alleviate such problems.

In another case a respondent was questioned by her management and the case discussed at the highest level in her trust as the manager of another HCP had raised the withdrawal as a safeguarding issue. The respondent found this a distressing experience, and though supported by the clinician involved in the withdrawal and her manager, still felt legally vulnerable.

In another case the professional disagreement about withdrawal caused a case of requested ventilation withdrawal to be taken through formal legal proceedings and to court. The case was upheld as legal withdrawal of treatment, but caused considerable distress to the patient’s family and considerable delay to the withdrawal.

### Emotional responses

The respondents in the doctors study indicated that though clear about the ethics and legality of withdrawal of assisted ventilation, most had mixed feeling about actually carrying it out and found the experience to be emotionally challenging.^13^ All but one of the HCPs in this sample felt that they were supporting the patient’s final wishes in assisting with withdrawal and achieving a peaceful death for a patient for whom treatment was futile, or their quality of life unbearable. This led to a great feeling of having done a good job. HCPs wanted to support patients and their families through these difficult decisions and processes and often felt privileged and honoured to be involved.

Despite the overall feeling of assisting a patient in ventilation withdrawal being the right thing to do, respondents still found the process emotionally and morally challenging.

> *HCP17, Morally and ethically, I think it was absolutely the right thing and I’m really chuffed we somehow pulled it out of the bag and did it so quickly …*

Withdrawing ventilation from patients with MND/ALS was described as ‘unusual’ and ‘odd’ and left HCPs and family members with ethical and moral questions. Many of our respondents felt that the process made them feel uncomfortable, or even scared. Some did not want the responsibility of removing ventilation and found the prospect ‘alarming’. Fear arose due to concern about symptom control being adequate to ensure a peaceful death, and where this was not achieved left the HCP distressed. For some HCPs who were supposed to be involved in a withdrawal, but in the end were not, there could be a sense of relief. For some it was setting a date for withdrawal which made them feel uncomfortable, as it felt like setting a time for someone to die depended on the HCPs who would be present. Some respondents were left wondering how long the patient may have lived for had they not chosen to remove ventilation. If this was perceived to be longer than a few weeks then HCPs could be left feeling troubled.

Among the family members there were perhaps fewer ethical and moral misgivings, commonly expressed as relief that the patient’s final wishes had been achieved. There were still mixed emotions and a large ethical burden for the relatives involved, which was made more difficult if delays in withdrawal had led to the patient’s loss of capacity. Many family members had very difficult discussions with patients about their wish to die and at what stage they would want to withdraw from treatment. Families and patients often discussed assisted suicide and euthanasia. Patients spoke to their family of their fears of death, or of dying without symptom relief, and also of their fears of being locked in. The primary reason for family members feeling these discussions to be necessary was the overriding desire to know what the patient’s wishes were so that they could ensure they were carried out. In most cases the family member felt easier about the withdrawal knowing that it was what the patient wanted, this made it morally correct.

> *R15 We talked endlessly about the ethics of it and the morality of it and that it’s…I think we had to be very clear in our minds that it’s okay to withdraw NIV, that it’s perfectly ethical, it isn’t assisted suicide. We had to get our heads round it*.

Results from our study seem to illustrate a correlation between the patient’s perceived closeness to death and the emotional acceptability of ventilation withdrawal. For many respondents, if the patient was perceived as being within days or weeks of death, then this made the ventilation withdrawal less ethically and legally challenging.

Where a patient was perceived to be alive, alert, mobile, and able to communicate, then this contributed towards feelings of uneasiness about the withdrawal and the perception that it was assisted death, or at least emotionally more challenging. Some participants felt that this perception of a patient as not being close to death was false and based on ignorance of mode of death in MND/ALS.

> *HCP18 … I think that all of the cases of withdrawal, you know whether it be an in-patient or at home, I’ve always felt that it was an end of life situation – I’ve never had an experience where I didn’t feel it should happen, I felt the ventilator was prolonging death and the dying process. (Palliative specialist)*

## DISCUSSION

Our research is the first to explore the experiences of families, nurses and allied health professionals involved in the withdrawal of ventilation at the request of a patient with MND/ALS. From the responses in this study it is clear that the experience of the withdrawal of assisted ventilation at the request of a patient with MND/ALS was a stressful and difficult experience for all involved. There appeared to be a lack of clarity, as to the right of the patient to stop treatment, even though the NICE guidance on NIV in MND/ALS has stressed the discussion of these aspects of future care.^3,8^ The right of a patient who has capacity for the decision to refuse treatment and for symptoms of withdrawal to be managed as required is clear in case law,^17,18^ and in the medical code of conduct in the UK.^19^

There is a need to ensure that this right to refuse or stop using an intervention and have symptoms managed in an alternative way is explained not only when NIV is commenced, when emotions may be profound, but throughout the disease progression and particularly when NIV is being used in the day time and near to 24 hours a day.^3,4^ All professional involved need to be more aware of the need for this discussion and the ethical and legal position. This may require complex and sensitive communication and reinforces the need for all professionals to receive communication skills training. This has been stressed in European guidelines for the care of patients with MND/ALS.^20,21^

Discussions with patients and families and within teams are underpinned by complex ethical dilemmas. Doctors have reported similar dilemmas in making these decisions and identified a lack of shared understanding as to the ethical, and legal, position.^12,13^ HCPs feared that their involvement in helping a person withdraw from assisted ventilation could be seen as abetting assisted suicide or assisted death. The intentions of the patient may be seen by some as “wanting to die”, rather than “being allowed to die”. However the ethical situation should be clear that the withdrawal of a treatment at the request of a patient with capacity for that decision is ethically sound and compliant with UK law.^22^ The challenge is to ensure that there is discussion with patient including assessment of the capacity for the decision and that the decision is settled and consistent and alternatives are known about and refused; discussion with the family and amongst the wider multidisciplinary team, so all involved are clear as to this position. National guidance was felt to be helpful in facilitating best outcomes for all parties.

Several of the respondents in this study had been faced with issues within their team, with disagreement or refusal to take part, or delays due to discussions about the ethical or legal aspects. This had added to the stress of the withdrawal for HCPs and patients and their families. HCPs in this study and doctors in a previous study talked of the time and emotional strain in ensuring that everyone did understand their role and the ethical issues.^12,13^ Families need reassurance that withdrawal is legal and ethical so they are not left with guilt and doubt.

The professionals involved saw their role as supporting families and carers, for which they needed correct legal and ethical knowledge. Lack of knowledge and experience was particularly evident among community nurses in this study and GPs in our previous study. Lack of knowledge and experience can lead to other professionals unknown to the patient and family getting involved at the last minute; patients not dying in their preferred place; delays in withdrawal; family members removing ventilation; or even inadequate symptom control during withdrawal. None of these scenarios is ideal patient care.

There were concerns expressed about finding the correct balance of the involvement for the family – not wanting them to feel responsible for the actions but at the same time not being excluded. Several respondents talked of the need to protect families from some of the stresses of the procedure, although they could not take away the overall distress of the death itself. This again did lead to uncertainty of how much to involve and include family members in the procedure and sensitive communication is necessary to ensure that the professionals are able to allow involvement, such as removal of the mask if they wish to do so, but not expecting them, to take an active part. It was felt that the professionals should take the lead and present a calm and confident face to patient and family, even if they were actually anxious and uncertain.

The respondents in the study interviewing doctors indicated that though clear about the ethics and legality of withdrawal of assisted ventilation, most had mixed feeling about actually carrying it out and found the experience to be emotionally challenging.^12^ These feeling were echoed in this study by both HCPs and family members. Withdrawal experience of the professionals was felt to be very important and all expressed the need for the involvement and / or advice of professionals who had been previous experience in withdrawal of assisted ventilation, to ease some of these concerns. Team support before, during and after the procedure was felt to be as important as that for families and carers. This support is often minimised within health and social care professions, although there is evidence of the need and effectiveness of support.^12,13^

The findings of this study have led to the development of Guidance for professionals by the Association for Palliative Medicine of Great Britain and Ireland (APM).^14, 22^ The Guidance is endorsed by several organisations including the MND Association, the Royal College of Physicians of London and the Royal College of Nursing in the UK. The APM also holds a list of volunteer mentors, clinicians with experience of this area of care who are willing to support others in withdrawing assisted ventilation at the request of a patient. The Guidance also calls for ongoing collection of a core dataset to allow continued quality assurance and outcomes evaluation.

## CONCLUSIONS

Health care professionals and relatives see the withdrawal of ventilation at the request of patients with MND/ALS as a difficult issue with ethical and legal challenges. There is a need for better understanding of the ethics and law so that patients, families and professionals feel supported and able to make clear decisions. Previous experience of withdrawal is very helpful and the involvement of experienced professionals with the whole multidisciplinary team is both supportive and reduces the burden of uncertainty and the impact of emotions on practice. There is a need for ethical and legal guidance so that all are clear before, during and after the withdrawal.

## Data Availability

All data produced in the present study are available upon reasonable request to the authors and with appropriate ethical permission. Participants were not asked for their consent for data to be used in other research.

